# Serological surveillance of SARS-CoV-2: Six-month trends and antibody response in a cohort of public health workers

**DOI:** 10.1101/2020.10.21.20216689

**Authors:** Ross J Harris, Heather J Whitaker, Nick J Andrews, Felicity Aiano, Zahin Amin-Chowdhury, Jessica Flood, Ray Borrow, Ezra Linley, Shazaad Ahmad, Lorraine Stapley, Bassam Hallis, Gayatri Amirthalingam, Katja Höschler, Ben Parker, Alex Horsley, Timothy J G Brooks, Kevin E Brown, Mary E Ramsay, Shamez N Ladhani

## Abstract

**Background:** Antibody waning after SARS-CoV-2 infection may result in reduction in long-term immunity following natural infection and vaccination, and is therefore a major public health issue. We undertook prospective serosurveillance in a large cohort of healthy adults from the start of the epidemic in England.

**Methods:** Clinical and non-clinical healthcare workers were recruited across three English regions and tested monthly from March to November 2020 for SARS-CoV-2 spike (S) protein and nucleoprotein (N) antibodies using five different immunoassays. In positive individuals, antibody responses and long-term trends were modelled using mixed effects regression.

**Findings:** In total, 2246 individuals attended 12,247 visits and 264 were seropositive in ≥2 assays. Most seroconversions occurred between March and April 2020. The assays showed >85% agreement for ever-positivity, although this changed markedly over time. Antibodies were detected earlier with Abbott (N) but declined rapidly thereafter. With the EuroImmun (S) and receptor-binding domain (RBD) assays, responses increased for 4 weeks then fell until week 12-16 before stabilising. For Roche (N), responses increased until 8 weeks, stabilised, then declined, but most remained above the positive threshold. For Roche (S), responses continued to climb over the full 24 weeks, with no sero-reversions. Predicted proportions sero-reverting after 52 weeks were 100% for Abbott, 59% (95% credible interval 50-68%) Euroimmun, 41% (30-52%) RBD, 10% (8-14%) Roche (N) <2% Roche (S).

**Interpretation:** Trends in SARS-CoV-2 antibodies following infection are highly dependent on the assay used. Ongoing serosurveillance using multiple assays is critical for monitoring the course and long-term progression of SARS-CoV-2 antibodies.

## Introduction

SARS-CoV-2 infection may be asymptomatic,(1) or manifest along a wide clinical spectrum, from mild upper respiratory tract illness to severe pneumonia, multiorgan failure and death.(2,3) Risk factors for COVID-19 include age, male gender, ethnicity, underlying comorbidity and occupation, especially being a healthcare worker.(4)

SARS-CoV-2 infection is usually confirmed by identifying viral RNA by RT-PCR of a nasal, nasopharyngeal or throat swab, but the sensitivity of this test may be as low as 70%,(5) which will significantly underestimate the true extent of infection in a population.(6) Antibody tests potentially provide a more accurate assessment of SARS-CoV-2 exposure and are a useful tool for understanding transmission dynamics and pandemic progression.(7,8) Following infection with COVID-19, antibodies are generated against a number of structural and non-structural proteins, including nucleoprotein (N) and spike protein (S). Antibodies against the receptor binding domain (RBD) of the spike protein block the interaction between the virus and the major host-cell receptor ACE2 and are likely to give the best correlation with neutralisation, and may protect against re-infection.(9,10) Until widespread coverage of an effective vaccine is achieved, COVID-19 control is likely to rely, at least in part, on herd immunity arising from a proportion of the population being exposed to the virus and developing a protective antibody response.(11) There are, however, concerns that SARS-CoV-2 antibodies may show significant waning, and antibody protection may be short-lived.(12–15)

In England, the first imported cases of COVID-19 were identified in late January 2020 and started increasing rapidly from early March, resulting in implementation of national lockdown on 23^rd^ March, which included closures of schools and non-essential businesses.(16) Cases continued to increase until mid-April before plateauing and then declining to low levels by end May 2020, after which lockdown measures since eased gradually. Studies of adult blood donors in particular estimated a seroprevalence of 5-10% across England and 15% in London during June 2020.(6,17,18) SARS-CoV-2 infection rates remained low during early summer and started increasing week-by-week from mid-August until the end of November 2020.

In order to monitor seroprevalence and the course of antibodies during the pandemic, Public Health England (PHE) initiated a monthly seroprevalence study in March 2020 across three English regions. Participants included healthcare workers with direct patient contact, those with public-facing but non-clinical roles, and non-clinical office and laboratory workers, who all continued working throughout the lockdown period. Repeated monthly testing allowed measurement of antibody seroconversion and sero-reversion rates as the pandemic unfolded, thus providing a unique opportunity to study SARS-CoV-2 antibody responses in a large cohort of healthy adults with asymptomatic infection and mild-to-moderate disease.

## Methods

### Study design

ESCAPE (**E**nhanced **S**eroIncidence for COVID-19 **A**ntibodies among **P**H**E** and NHS Staff) is a prospective surveillance cohort collecting blood samples for SARS-CoV-2 antibodies and questionnaire data at monthly intervals. Surveillance was initiated in March 2020 and included staff from Public Health England (PHE), National Health Service (NHS) and volunteers across four sites. Sites were (1) London (PHE London and Colindale, primarily office and laboratory staff), (2) PHE Porton Down (Southwest England, primarily office and laboratory staff in rural Wiltshire plus family and friends), (3) PHE Manchester and Manchester Royal Infirmary (MRI) (office and laboratory staff as well as healthcare workers) and (4) NHS Wythenshawe Hospital, Manchester (primarily frontline healthcare workers). After providing written consent, participants completed a brief questionnaire about recent respiratory symptoms and provided ∼10 mL of blood by venepuncture. Samples were processed in PHE laboratories and frozen at −70°C or below until they were batch-tested for SARS-CoV-2 antibodies. Participants who missed an appointment continued to be invited for subsequent appointments, as did participants who became unwell with a respiratory or any other illness.

### Laboratory methods

Samples were tested with four different commercial serological assays and an in-house assay, against N and S antigens in two broad assay types: an indirect IgG format and a double-antigen total antibody format. Specifically,

1. **EUROIMMUN:** an enzyme-linked immunosorbent assay (ELISA) for semi-quantitative detection of IgG antibodies using recombinant S1 domain of SARS-CoV-2 spike protein (EUROIMMUN Medizinische Labordiagnostika AG, Lubeck, Germany). Results were reported as an index relative to the optical difference of a reference sample. Indices >1.1 were considered positive. Results plateau with indices >15.
2. **Roche S:** The Elecsys Anti-SARS-CoV-2 S assay is an electrochemiluminescant immunoassay that detects total high-affinity antibody (IgG and IgM) against RBD (Roche Diagnostics International Ltd, Rotkreuz, Switzerland). Results are provided against a standard curve and reported as U/mL. A result of ≥0.8 is considered positive.
3. **Roche N:** Similar to Roche S, this assay detects IgG and IgM against nucleoprotein. The results are provided as an index against a reference sample. An index of ≥1.0 is considered positive.
4. **Abbott:** SARS-CoV-2 IgG assay is a chemiluminescence enzyme immunoassay for qualitative detection of IgG antibodies against nucleoprotein (Abbott Diagnostics, IL, USA). The assay using a bead-based indirect format with ‘blockers’ to decrease cross-reaction with antibodies to other coronavirus N proteins.
5. **RBD:** The in-house RBD assay is an indirect IgG assay, developed and validated at PHE. Commercial RBD subunit was purchased from SinoBiological Inc (Beijing, P.R. China) and expressed in HEK293 cell culture with a C-terminal mouse Fc tag (Arg319-Phe541(V367F);# YP_009724390.1). Nunc MaxiSorp flat-bottomed, polystyrene 96-well microtitre plates were coated by diluting 20ng recombinant protein/well in sterile PBS; pH7.2 ± 0.05 (-CaCl2, - MgCl2), (GIBCO, Thermo Fischer, Waltham, U.S.A) at 4-8°C for a minimum of 16 hours. Serum was diluted at a final dilution factor of 1 in 100. IgG binding on the plate surface was detected with an anti-Human IgG−horseradish peroxidase antibody conjugate (Sigma Aldrich, St Louis, U.S.A) and detected with 3,3⍰,5,5⍰-Tetramethylbenzidine (Europa Bioproducts Ltd, Ipswich, U.K). Samples were analysed in duplicate and optical density (OD_450_) data were evaluated by dividing average OD_450_ values for individual samples by average OD_450_ of a known calibrator with negative antibody levels (T/N ratio). Results ≥5 were considered positive.

For all assays, specificity and sensitivity were determined using a panel of negative samples collected in 2019, and of confirmed PCR positive samples collected at the beginning of the outbreak, respectively.

### Statistical analysis

#### Seroprevalence, conversions and reversions

All commercial assays were run as singletons, the in-house RBD assay was run as duplicate. Data for each assay were analysed separately. Because only a proportion of samples were tested with the RBD assay, seroprevalence was not calculated for this assay. The proportion positive at each visit for the four sites was calculated with 95% binomial confidence intervals (CI). Seroconversion was defined as having a positive test subsequent to a negative test at the previous visit. Sero-*reversion* was defined as having a negative test subsequent to a positive test at the previous visit. The proportion of seroconversions and reversions was calculated with 95% confidence intervals (CI) across study visits. We also calculated the proportion ever positive throughout the study.

Agreement in positivity under each pair of assays was assessed using the kappa statistic, where 0 indicates no agreement and 1 perfect agreement. We examined agreement between tests according to calendar month, and ever-positive status in participants. 95% CI for kappa statistics were obtained via bootstrap resampling, with the 2.5^th^ and 97.5^th^ percentiles of 1000 samples forming the lower and upper bounds.

#### Predictors of initial response

Individuals with at least one positive test under two different assays (EuroImmun, either of Roche N/S, and Abbott) were classified as “confirmed positive”. For each assay, the maximum level reached within 12 weeks of their first positive test (under any assay) was taken as the initial response. Linear regression models were fitted to the log of the measurements with the following covariates: age (18-29, 30-39, 40-54 and 55+), sex, ever-reported SARS-CoV-2 RT-PCR positive, respiratory symptoms (a) 0-28 days prior to their first positive antibody test, and (b) 29-84 days prior. Interactions between covariates were assessed via likelihood ratio tests.

#### Trends in antibody response post-12 weeks

Following the initial 12-week response in the confirmed seropositive cohort, we modelled the log of the measurements, with time as a continuous variable, from 12 weeks after first positive test. Random effects representing individual variability in response were included for the intercept (baseline result at 12 weeks) and slopes (change over time). Correlation between the random effect for the intercept and slope was allowed for in the variance-covariance matrix. The same covariates were considered as predictors for baseline result (week 12) and change over time, the latter as covariate X time interactions. Data were analysed using Stata v14.2 (StataCorp, College Station, Texas).

#### Joint modelling and predictions

The random effects model for trends post-12 weeks was also implemented in a fully Bayesian Markov Chain Monte Carlo (MCMC) framework; this model only included random intercepts and trends over time. Results for all assays were modelled simultaneously, allowing an assessment of individual-level correlations between trends for different assays. The model was used to derive predicted responses up to 52 weeks, based on the estimated individual-level random effects. From these, predicted positivity and multivariate assay status was obtained. The MCMC approach allows uncertainty of such derived results to be quantified, with the median of the posterior distribution taken to be the point estimate, and the 2.5^th^ and 97.5^th^ percentiles forming a 95% credible interval (CrI). The model was fitted using WinBUGS 1.4.3 (MRC Biostatistics Unit).

## Results

### Summary statistics

In total, 2246 individuals were recruited (London, 537; Manchester, 594; Wythenshawe, 598 and Porton, 517) and 2045 (91%) had at least 3 visits, with up to 8 visits (median=6), between 23^rd^ March and 18^th^ November 2020. 1,101 (49%) were NHS employees. The median interval between study visits was 28 days (90% of visits between 23-56 days). There were 12,247 attendances and 95.2% of scheduled visits were attended. The median age of study participants at entry was 40 years (interquartile range 32-50, range 18-71) and 69% (n=1529) were female. Only 52 individuals (2.3%) reported having a lab-confirmed COVID-19 diagnosis, while 580 (26%) reported having a respiratory illness.

### Seroprevalence, conversions and reversions

There were 12,230 EuroImmun tests, 11,265 Roche N, 10,980 Roche S, 9,569 Abbott and 3,694 RBD tests completed. Ever-positivity in study participants was 260/2246 (11.6%) for EuroImmun, 257/2245 (11.5%) for Roche N, 264/2245 (11.8%) for Roche S, and 279/2212 (12.6%) for Abbott. For RBD, a higher proportion (270/1141, 15.8%) tested positive because of selective testing of individuals positive for other assays.

Ever-positivity using EuroImmun was highest among Wythenshawe hospital employees (22.2%) followed by PHE London (13.6%), Manchester (7.4%) and PHE Porton (1.8%). Positivity was 14.7% in those aged 15-29, compared to 30-39: 11.5%, 40-54: 10.7%, ≥55: 9.1%. There was no difference between males and females (chi-square p-value=0.639). Positivity was 55.8% in those reporting laboratory-confirmed COVID-19 (10.4% in those without) and 25.4% in those reporting a respiratory illness (6.6% in those without). Similar patterns were observed with the other assays.

Seropositivity was lowest in March and increased in April at all sites (Figure 1, Supplementary Table S1). This is also reflected in the seroconversion rates which ranged from 6.1-6.8% across the four assays in April, 3.5-4.2% in May, 0.9-1.2% in June and <0.6% in July. Few seroconversions were observed subsequently and none in November, although data are sparse. Conversely, sero-reversions increased over time, reaching 10%/month for EuroImmun and 10.5% for Abbott in July, and 18.5% for EuroImmun and 33% for Abbott in October.

**Figure 1:**
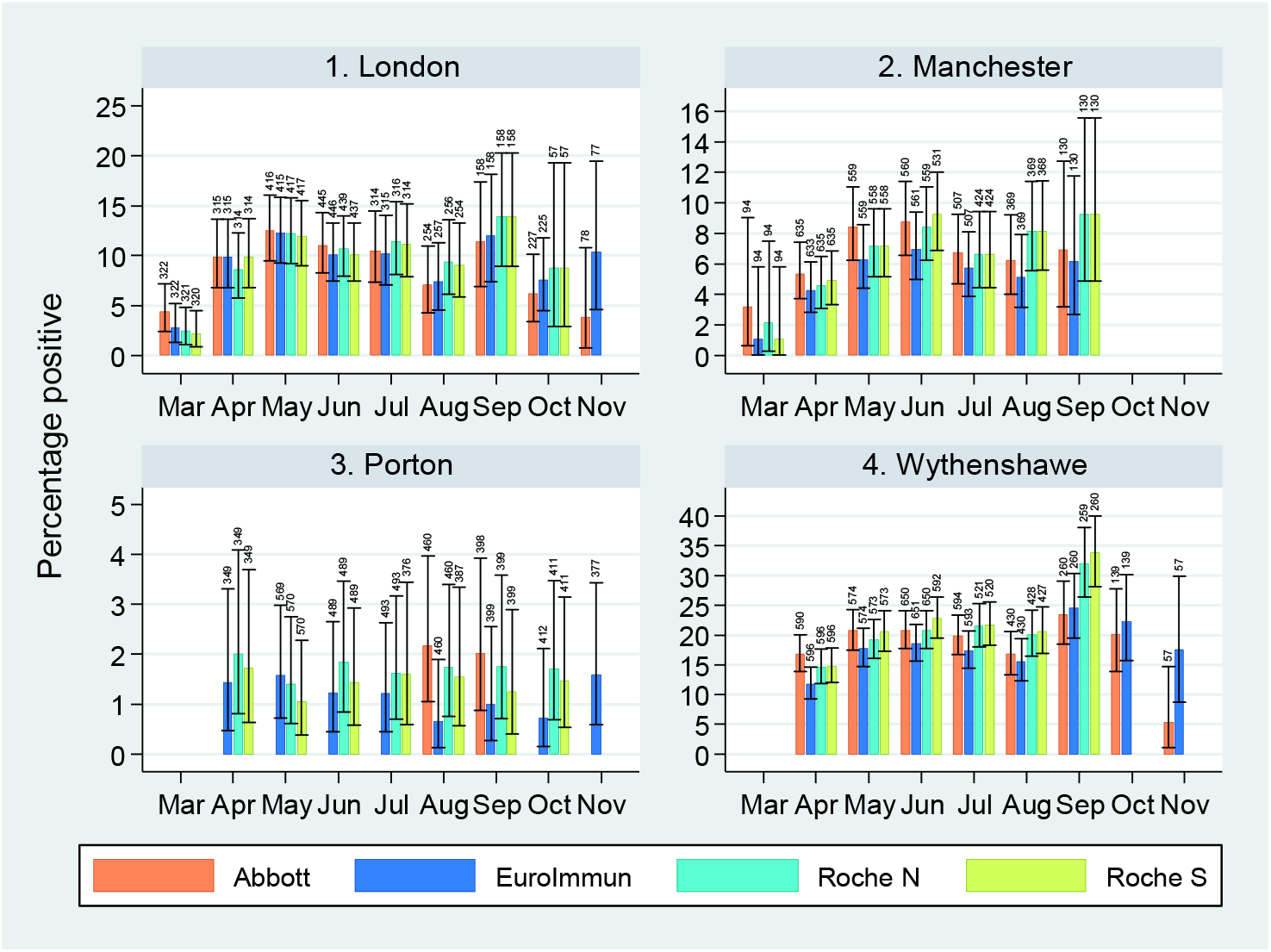
% positive (spike protein IgG) or % positive/indeterminate (nucleoprotein IgG), by assay, study site and calendar month, with binomial 95% confidence intervals. Sample sizes are indicated above each bar. **Note:** There were 21 Roche N/S tests for Wythenshawe in October with majority positive, which are not shown. The high observed prevalence is likely due to sampling of particular individuals: 260 were sampled in September.

### Antibody trends

There were 264 individuals testing positive in ≥2 assays (i.e. EuroImmun, Roche N or S, and Abbott), considered “confirmed positives”. For Abbott, EuroImmun and RBD, the initial antibody response was rapid following their last negative test (Figure 2). Levels then drop immediately for Abbott and continue to fall over the following 28 weeks, with a high proportion sero-reverting. With EuroImmun, responses increased for 4 weeks after first positive test, then fell to week 12-16 before stabilising, although a proportion sero-reverted. Similar patterns were observed for RBD, although responses were higher above the positive threshold and fewer sero-reverted. The Roche assays behaved differently: for Roche N, responses climbed until 8 weeks, stabilised, and began to fall, but the majority were well above the positive threshold. For the Roche S assay, responses continued to climb over the full 24 weeks, with no sero-reversions.

**Figure 2.**
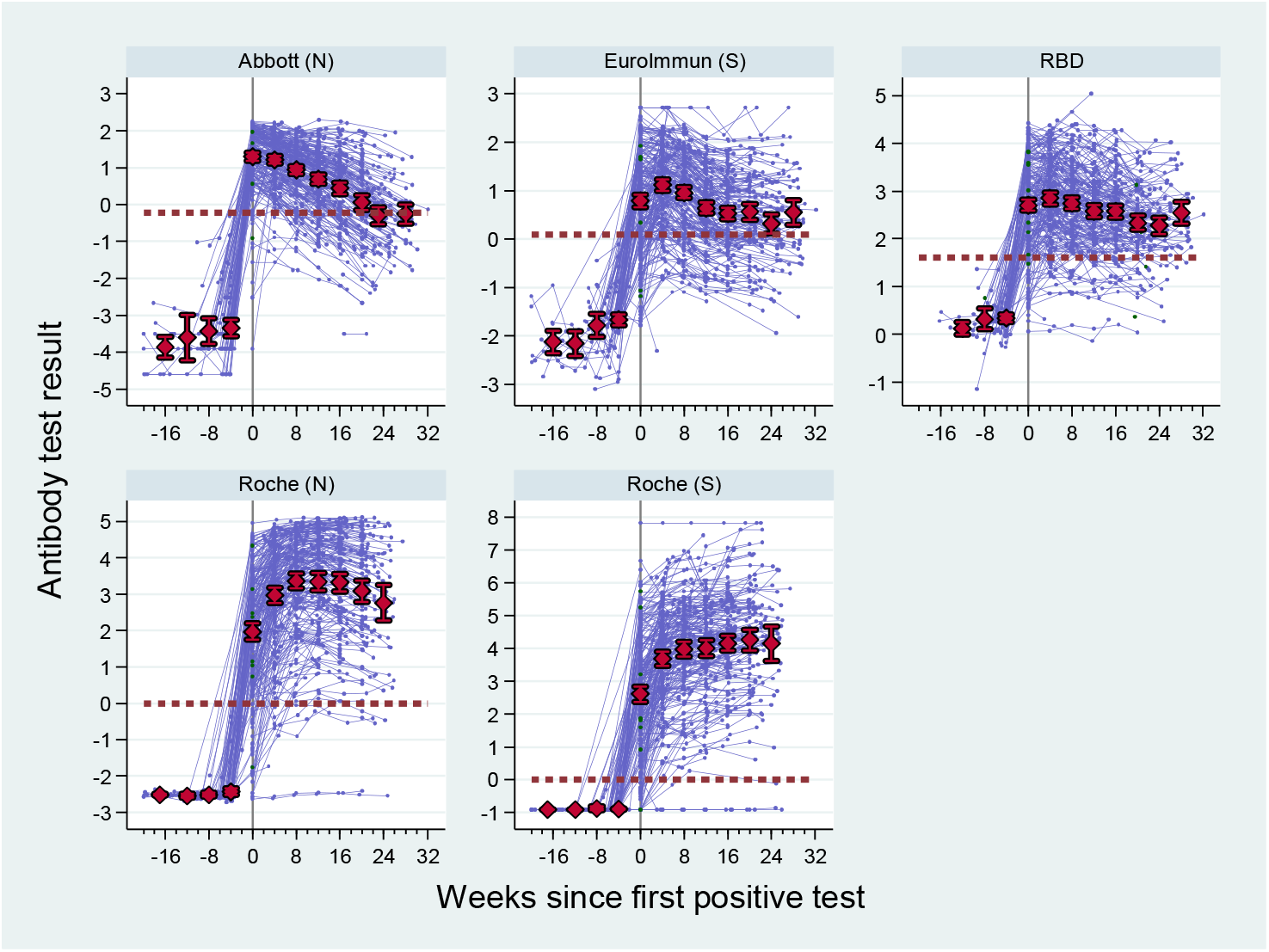
Trends in antibody response in confirmed positives (at least two of EuroImmun, Roche N and Abbott). Means and bars for 95% confidence intervals are superimposed on the plot. Reference lines on the y-axis indicate positive threshold for the tests, the reference line at week 0 indicates time of first positive test (any assay).

Figure 3 shows kappa statistics for agreement between the assays. All assays showed >85% agreement for ever-positivity, although this changed markedly over time, with poor agreement in March, rising to a peak in May and then declining, with the exception of Roche N/Roche S, and particularly poor agreement for Abbott. This reflects the longer interval after infection in latter months, and the large proportion reverting to sero-negativity for EuroImmun and Abbott.

**Figure 3.**
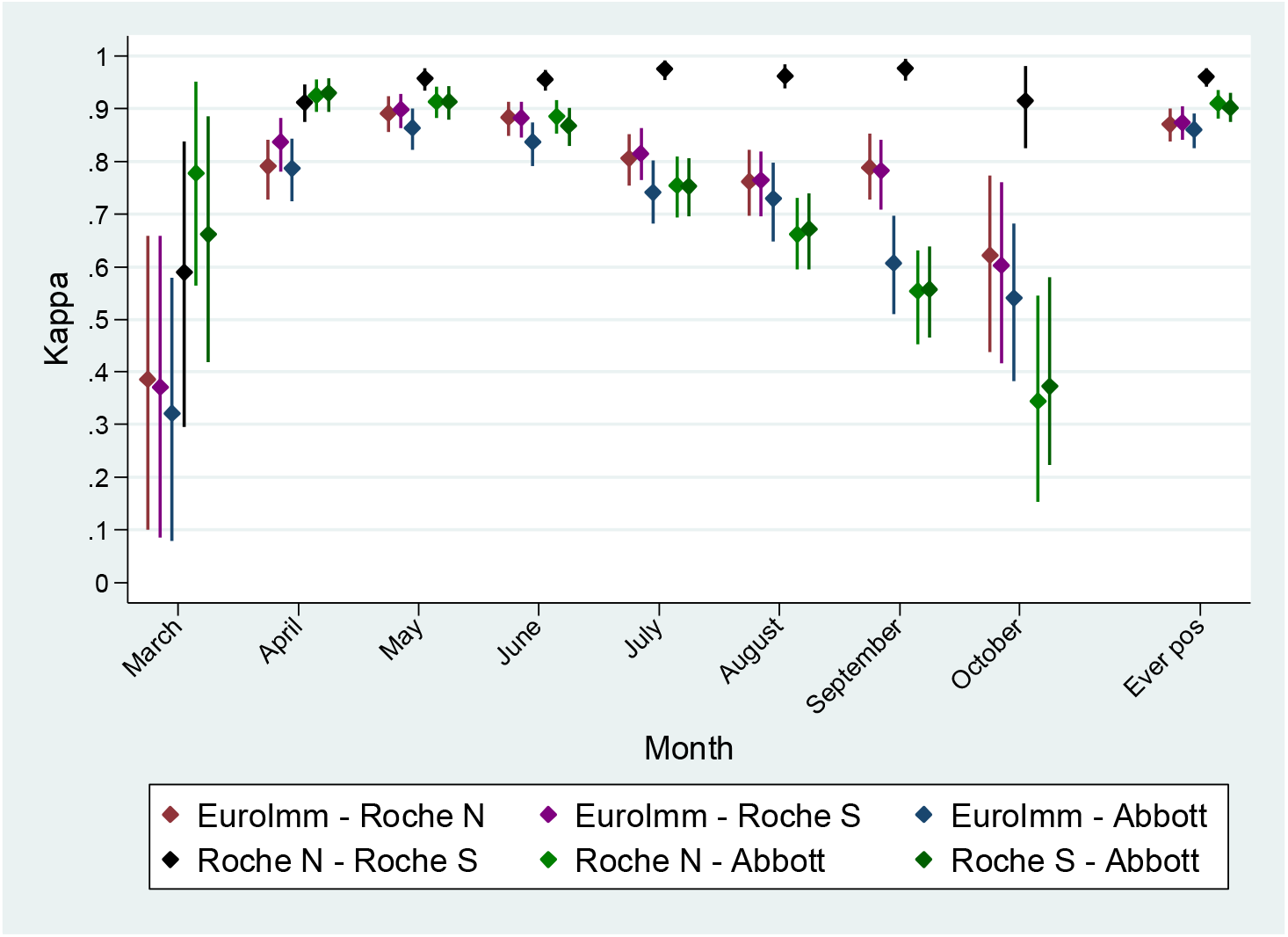
Kappa statistics with 95% confidence intervals for agreement between assays over the study period.

### Initial antibody response

For all assays, the regression model for maximum antibody 12 weeks after first positive test found no difference by age (minimum LR test p-value=0.393) or sex (minimum LR test p-value=0.604). Those with confirmed COVID-19 had slightly higher antibody levels for EuroImmun (ratio, 1.45; 95%CI. 1.03-2.03) but not for other assays (minimum p-value=0.226), although confidence intervals are wide. Those reporting respiratory symptoms up to 28 days before first seropositivity had higher responses under all assays, with 29% (EuroImmun) to 56% (Roche S) higher ratios and moderate-to-strong evidence for all assays. There was no difference in response for those reporting a respiratory illness 29-84 days prior to first seropositivity (minimum p-value=0.399).

Tests for interactions between age, sex, respiratory symptoms and COVID-19 diagnosis showed little evidence for interactions. Older individuals with a COVID-19 diagnosis had higher measurements using the two Roche assays (LR test for interaction p-value =0.003 for Roche N, p=0.007 for Roche S); this was not observed for the other assays. Notably, only 28 seropositive individuals reported a PCR-confirmed COVID-19 diagnosis. Of the 52 individuals in the whole study reporting confirmed COVID-19, 22 (42%) tested positive for all five assays, 6 for ≥2assays, 4 for one assay and 20 (38%) remained seronegative for all assays, despite attending multiple (4-8) visits.

### Trends in antibody response post-12 weeks

The random effects model for antibody response 12 weeks after first seropositivity was initially adjusted for age, sex, confirmed COVID-19, and respiratory symptoms within 28 days of seropositivity. Weekly changes in antibody responses were: −1.6% (−2.14% to −1.18%) for EuroImmun, −3.54% (−4.32% to −2.75%) for Roche N, 2.60% (1.60% to 3.62%) for Roche S (an increase), −7.85% (−8.38% to −7.32%) for Abbott, and −1.82% (−2.41% to −1.23%) for RBD. Covariates were then included as predictors of trends (Table 3). Age, sex, confirmed COVID-19 and respiratory symptoms were not predictive of trends from 12 weeks onwards, except for Roche S, which showed a steeper increasing trend in ≥55 year-olds compared to other age-groups (11.4%/ week). Increases over time were also greater in those with confirmed COVID-19, although data for this group were relatively sparse.

The variance components indicated substantial individual variability in base response, but comparatively little variation in trends, which were generally stable (Supplementary Figure S2). For most assays, there was little correlation between baseline response and trend, but for Abbott there was some positive correlation (higher initial responses were associated with slower declines).

### Joint modelling and predictions

The Bayesian model for trends allows investigation of the correlation between individual responses under different assays (Supplementary Table S3). Correlation between baseline responses (random effects for the intercept) ranged from 0.399 for Roche S/Abbott to 0.785 for EuroImmun/RBD. Correlations between intercepts and trends were generally weak. As in the classical model, there was modest positive correlation (0.113) between the intercept and slope for RBD, and very little between-assay correlation (i.e., baseline result for one assay predicting the trend of another assay). There were modest correlations in trends between some assays, the strongest being EuroImmun and Roche S (0.249), and Abbott and Roche N (0.238); and weaker for EuroImmun/Roche N (0.147) and Roche N/Roche S (0.158).

By 26 weeks, 37% (95%CrI, 34-40%) of seropositive individuals were predicted to be seronegative for EuroImmun, 5% (4-6%) for Roche N, 2% (1-2%) for Roche S, 49% (45-53%) for Abbott and 16% (13-19%) for RBD (Supplement Figure S4). Predicted proportions sero-reverting continued to increase for EuroImmune, with 59% (95% CrI 50-68%) seronegative at 52 weeks. Sero-reversion at 52 weeks were lower for the Roche N (10% (8-14%) and Roche S (<2%) assays. For RBD, the predicted proportion negative accelerated over time, reaching 41% (30-52%) at 52 weeks. Sero-reversion increased most rapidly with the Abbott assay with all individuals sero-reverting by week 45.

Supplementary figure S5 shows predicted multivariate assay status. At 13 weeks after first seropositivity, 64% are expected to be positive for all assays, but this drops to <10% after 36 weeks due to rapid waning with the Abbott assay. By 52 weeks, <40% remained positive for EuroImmun, RBD and both Roche assays, 19% remained positive for RBD and both Roche assays, and 31% for both Roche assays only. Overall, 8% were predicted to be negative at 52 weeks for all assays except Roche S, which was predicted to remain positive for almost all individuals throughout, given the linear trend.

## Discussion

We analysed data from a monthly prospective longitudinal cohort study of 2247 PHE and NHS employees in three English regions, with up to eight sampling time-points from the start of the COVID-19 epidemic in March 2020 until mid-November 2020. Seroprevalence by region and time was consistent with other sources, with a higher seroprevalence in London, a rise in seroprevalence from March to May 2020 and a decline from June to August.(6) Healthcare workers with direct patient contact had nearly double the seroprevalence of PHE London staff, which are not in patient-facing roles. (4) Almost all infections occurred early in the surveillance, with low seroconversion rates during summer and a small increase in October detected by the Abbott assay, likely because of a slower response for the other assays. Overall agreement between assays was >85% for ever-positivity, with similar proportions ever-positive across assays. Agreement between individual tests, however, declined with calendar time, as participants in the study reverted to negativity for some assays but not others.

Antibodies were detected earliest with the Abbott (nucleoprotein) assay which is advantageous for confirming SARS-CoV-2 infection rapidly after virus exposure, but declined rapidly, with all predicted to revert to negative after one year. Responses were still rising one month after first positive test for EuroImmun (spike protein) and RBD, before falling until week 12 and then stabilising. Despite low declining rates beyond 12 weeks, 59% were predicted to sero-revert by 52 weeks for EuroImmun and 41% for RBD. Responses for the Roche assays rose most slowly, with Roche N starting to decline from 12 weeks but remaining well above the positive threshold, and the majority likely to remain positive if trends continued. At 6 months, there was no evidence of a decline for Roche S, although an eventual decline seems likely. Further studies are needed to understand the rapid decline in seropositivity over time with the Abbott assay and the lack of decline with the Roche S assay.

The strength of this surveillance was the early recruitment of large numbers of seronegative participants at the start of the epidemic and high follow-up rates. We recruited both clinical and non-clinical staff in three English regions, most of whom continued to work during national lockdown. The monthly follow-up with blood sampling allowed longitudinal assessment of seroprevalence against multiple viral antigens in a large population of healthy adults who were exposed to SARS-CoV-2, most of whom were asymptomatic or had mild-to-moderate disease.

Following infection with other seasonal coronaviruses, anti-nucleocapsid antibodies decline within two months and reinfections are common after 12 months. (19) Due to the relative novelty of SARS-CoV-2, longitudinal antibody studies so far have primarily included small numbers of mainly hospitalised patients with severe COVID-19 with limited follow-up.(13,20–23). One such study suggested more rapid waning of antibodies, with 12/30 (40.0%) asymptomatic individuals and 4/31 (12.9%) symptomatic individuals sero-reverting within 8 weeks of hospital discharge.(24) Another report involving 34 patients followed-up for a mean of 86 days after infection estimated a half-life of 36 days for SARS-CoV-2 antibodies.(25) Short-lasting immunity would suggest that SARS-CoV-2 could enter into regular circulation alongside other coronaviruses, with seasonal variation depending on the length of protection from the initial infection.(15) Our findings, however, which include over 2000 healthy adults and, therefore, more representative of the general population, indicate that SARS-CoV-2 antibodies do not decline as quickly as predicted by smaller cohorts of patients with shorter follow-up. (13,20–23) Our findings are consistent with the recent Icelandic study which found most hospitalised patients with COVID-19 remained seropositive 120 days after diagnosis with no significant decrease in antibody levels using two different antibody assays.(26)

The humoral response is considered to provide the first line of defence against infection and, therefore, the presence, neutralising ability and persistence of antibodies is likely to correlate with protection against infection,(27) which is consistent with our understanding of the host immune response to other respiratory viruses. SARS-CoV-2 infection also triggers a cellular immune response, with activation of a range of T cells against all major SARS-CoV-2 antigens, which is independent to B-cell mediated antibody responses.(28,29) The strong T cell responses after SARS-CoV-2 infection would support long-term protection even in the absence of detectable antibodies.(30,31)

For both MERS-CoV and SARS-CoV, antibodies were shown to last for at least two years after infection.(32,33) The duration of immunity following SARS-CoV-2 infection is a critical factor in determining the course of the pandemic.(15) Ongoing serosurveillance will remain critical for monitoring the course and projection of SARS-CoV-2 antibodies in the longer term.

## Supporting information

Table S1. Full seroprevalence results by site over time for 4 assays

## Data Availability

Sharing of individual-level data is not permitted under the study protocol, although provision of aggregated data for specific purposes will be considered on request.

## Ethics Approval

This study was approved by PHE Research Support and Governance Office (R&D REGG Ref NR 0190).

## Funding

This study was internally funded by Public Health England

## Author contributions

SL was the chief investigator, had full access to all the data in the study and had final responsibility for the decision to submit for publication. SL, MR, BH, SA, RB and NA conceived and designed the study. EL, TB and KB oversaw data collection and quality assurance. RH and HW analysed the data and carried out data checking. RH, HW and SL drafted the manuscript. All authors contributed to interpretation of the data and critical revision of the final manuscript.

## Acknowledgements

BP and AH are supported by the NIHR Manchester Biomedical Research Centre.

This research was internally funded by Public Health England and carried out at by the NIHR Manchester Clinical Research Facility. The views expressed are those of the authors and not necessarily those of Public Health England, NHS, the NIHR or the Department of Health.

We would like to express our gratitude to all the participants and to the laboratory staff for testing the samples.

---

**Table 1.** Predictors of initial response in confirmed positive individuals; respiratory symptoms variable defined as: 1) no symptoms 84 days before first positive test, 2) symptoms within 28 days before first positive test, and 3) symptoms 29-84 days before first positive test. Coefficients from the model are exponentiated to provide estimates as ratios in levels between groups.

|  | **Eurolmmun** |  | **Roche N** |  | **Roche S** |  | **Abbott** |  | **RBD** |  |
| --- | --- | --- | --- | --- | --- | --- | --- | --- | --- | --- |
|  | Estimate (95% CI) | p-val | Estimate (95% CI) | p-val | Estimate (95% CI) | p-val | Estimate (95% CI) | p-val | Estimate (95% CI) | p-val |
| Age 15-29 | 1.05 (0.79, 1.38) | 0.746 | 1.09 (0.60, 1.96) | 0.774 | 1.01 (0.55, 1.83) | 0.982 | 1.11 (0.87, 1.42) | 0.388 | 1.08 (0.81, 1.44) | 0.608 |
| Age 30-39 | 1 (base) |  | 1 (base) |  | 1 (base) |  | 1 (base) |  | 1 (base) |  |
| Age 40-54 | 1.07 (0.82, 1.40) | 0.615 | 0.96 (0.54, 1.68) | 0.873 | 0.99 (0.56, 1.75) | 0.961 | 1.10 (0.87, 1.39) | 0.417 | 1.15 (0.87, 1.53) | 0.311 |
| Age 55+ | 1.18 (0.84, 1.66) | 0.345 | 1.63 (0.78, 3.38) | 0.191 | 1.22 (0.59, 2.54) | 0.593 | 1.29 (0.96, 1.74) | 0.092 | 1.15 (0.81, 1.65) | 0.438 |
| Female vs. male | 0.94 (0.75, 1.18) | 0.610 | 0.92 (0.57, 1.48) | 0.733 | 0.98 (0.60, 1.58) | 0.920 | 1.00 (0.82, 1.22) | 0.988 | 1.07 (0.84, 1.35) | 0.590 |
| COVID diag vs. not | 1.45 (1.03, 2.03) | 0.033 | 0.81 (0.39, 1.65) | 0.558 | 1.53 (0.74, 3.15) | 0.252 | 0.96 (0.72, 1.29) | 0.789 | 1.25 (0.87, 1.80) | 0.226 |
| No symptoms | 1 (base) |  | 1 (base) |  | 1 (base) |  | 1 (base) |  | 1 (base) |  |
| Symp <=28 days | 1.28 (1.02, 1.61) | 0.033 | 1.44 (0.89, 2.34) | 0.137 | 1.56 (0.96, 2.54) | 0.073 | 1.24 (1.01, 1.51) | 0.035 | 1.37 (1.07, 1.74) | 0.011 |
| Symp 29-84 days | 1.07 (0.80, 1.44) | 0.627 | 1.08 (0.58, 2.00) | 0.805 | 1.10 (0.59, 2.05) | 0.772 | 1.11 (0.87, 1.43) | 0.399 | 1.00 (0.74, 1.36) | 0.981 |
| Constant | 2.88 (2.20, 3.76) | 0.000 | 19.8 (11.2, 34.9) | 0.000 | 34.6 (19.5, 61.4) | 0.000 | 3.42 (2.70, 4.35) | 0.000 | 15.7 (11.8, 20.8) | 0.000 |

**Table 2.** Predictors of baseline response and trends from 12 weeks after first positive test in confirmed positive individuals. Respiratory symptoms variable defined as symptoms within 28 days before first positive test. Coefficients from the model are exponentiated to provide estimates as ratios in levels between groups, or ratio changes per week for trends.

|  | Eurolmmun |  | Roche N |  | Roche S |  | Abbott |  | RBD |  |
| --- | --- | --- | --- | --- | --- | --- | --- | --- | --- | --- |
|  | Estimate (95% CI) | p-val | Estimate (95% CI) | p-val | Estimate (95% CI) | p-val | Estimate (95% CI) | p-val | Estimate (95% CI) | p-val |
| **Baseline response** |  |  |  |  |  |  |  |  |  |  |
| Age 15-29 | 1.07 (0.80, 1.44) | 0.635 | 1.16 (0.64, 2.10) | 0.626 | 0.98 (0.55, 1.75) | 0.957 | 1.23 (0.83, 1.81) | 0.303 | 1.10 (0.80, 1.51) | 0.546 |
| Age 30-39 | 1 (base) |  | 1 (base) |  | 1 (base) |  | 1 (base) |  | 1 (base) |  |
| Age 40-54 | 1.13 (0.86, 1.49) | 0.380 | 0.91 (0.53, 1.56) | 0.731 | 0.90 (0.53, 1.53) | 0.702 | 1.32 (0.92, 1.89) | 0.136 | 1.12 (0.84, 1.50) | 0.442 |
| Age 55+ | 1.27 (0.90, 1.79) | 0.169 | 1.48 (0.74, 2.93) | 0.266 | 1.10 (0.56, 2.15) | 0.779 | 1.61 (1.03, 2.53) | 0.037 | 1.38 (0.96, 1.98) | 0.084 |
| Female vs. male | 1.06 (0.84, 1.34) | 0.639 | 1.27 (0.79, 2.04) | 0.316 | 1.42 (0.90, 2.25) | 0.132 | 0.92 (0.70, 1.21) | 0.554 | 1.05 (0.82, 1.35) | 0.692 |
| COVID diag vs. not | 1.54 (1.05, 2.26) | 0.026 | 1.82 (0.84, 3.96) | 0.130 | 2.70 (1.27, 5.75) | 0.010 | 1.13 (0.72, 1.76) | 0.594 | 1.27 (0.83, 1.94) | 0.268 |
| Symp <=28 days | 1.32 (1.06, 1.65) | 0.013 | 1.20 (0.77, 1.87) | 0.414 | 1.33 (0.86, 2.04) | 0.197 | 1.21 (0.93, 1.56) | 0.158 | 1.21 (0.96, 1.53) | 0.102 |
| **Trend** |  |  |  |  |  |  |  |  |  |  |
| Base group trend | 0.977 (0.964, 0.990) | 0.001 | 0.977 (0.954, 1.000) | 0.051 | 1.000 (0.975, 1.026) | 0.994 | 0.934 (0.919, 0.950) | 0.000 | 0.976 (0.959, 0.993) | 0.005 |
| Age 15-29 | 0.987 (0.973, 1.001) | 0.066 | 0.998 (0.975, 1.022) | 0.878 | 0.990 (0.965, 1.016) | 0.461 | 0.999 (0.982, 1.016) | 0.886 | 0.997 (0.980, 1.014) | 0.720 |
| Age 30-39 | 1 (base) |  | 1 (base) |  | 1 (base) |  | 1 (base) |  | 1 (base) |  |
| Age 40-54 | 1.001 (0.989, 1.013) | 0.848 | 1.011 (0.990, 1.034) | 0.303 | 1.013 (0.989, 1.037) | 0.288 | 1.001 (0.986, 1.016) | 0.911 | 0.989 (0.974, 1.004) | 0.143 |
| Age 55+ | 1.009 (0.995, 1.023) | 0.226 | 1.017 (0.992, 1.043) | 0.182 | 1.058 (1.030, 1.087) | 0.000 | 1.001 (0.983, 1.019) | 0.932 | 0.993 (0.976, 1.011) | 0.427 |
| Female vs. male | 1.013 (1.002, 1.023) | 0.021 | 0.978 (0.960, 0.996) | 0.016 | 1.018 (0.998, 1.039) | 0.081 | 0.989 (0.976, 1.002) | 0.097 | 1.015 (1.001, 1.029) | 0.031 |

|  | EuroImmun |  | Roche N |  | Roche S |  | Abbott |  | RBD |  |
| --- | --- | --- | --- | --- | --- | --- | --- | --- | --- | --- |
|  | Estimate (95% CI) | p-val | Estimate (95% CI) | p-val | Estimate (95% CI) | p-val | Estimate (95% CI) | p-val | Estimate (95% CI) | p-val |
| COVID diag vs. not | 1.001 (0.985, 1.018) | 0.885 | 0.998 (0.968, 1.030) | 0.909 | 1.042 (1.007, 1.079) | 0.019 | 0.983 (0.963, 1.003) | 0.101 | 1.007 (0.985, 1.030) | 0.531 |
| Symp <=28 days | 0.994 (0.985, 1.004) | 0.248 | 0.992 (0.976, 1.009) | 0.370 | 0.993 (0.975, 1.011) | 0.426 | 0.989 (0.977, 1.001) | 0.063 | 1.001 (0.990, 1.013) | 0.847 |
| Constant | 1.49 (1.12, 1.98) | 0.006 | 25.79 (14.38, 46.27) | 0.000 | 37.15 (21.08, 65.48) | 0.000 | 1.95 (1.38, 2.74) | 0.000 | 10.97 (8.07, 14.90) | 0.000 |
| **Variances** |  |  |  |  |  |  |  |  |  |  |
| Trend | 0.0004<br>(0.0003, 0.0007) | 0.000 | 0.0017<br>(0.0012, 0.0024) | 0.000 | 0.0016<br>(0.0011, 0.0023) | 0.000 | 0.0009<br>(0.0006, 0.0012) | 0.000 | 0.0003<br>(0.0001, 0.0007) | 0.000 |
| Base response | 0.5291<br>(0.4287, 0.6532) | 0.000 | 2.0689<br>(1.6832, 2.5431) | 0.000 | 1.9322<br>(1.5649, 2.3856) | 0.000 | 0.7357<br>(0.5982, 0.9048) | 0.000 | 0.5536<br>(0.4415, 0.6941) | 0.000 |
| Trend X base covariance | 0.0022<br>(-0.0012, 0.0056) | 0.209 | -0.0041<br>(-0.0152, 0.0070) | 0.468 | 0.0023<br>(-0.0092, 0.0137) | 0.697 | 0.0161<br>(0.0105, 0.0216) | 0.000 | -0.0042<br>(-0.0091, 0.0008) | 0.100 |
| Residual | 0.0178<br>(0.0145, 0.0219) | 0.000 | 0.0082<br>(0.0062, 0.0107) | 0.000 | 0.0221<br>(0.0172, 0.0284) | 0.000 | 0.0164<br>(0.0132, 0.0203) | 0.000 | 0.0471<br>(0.0385, 0.0577) | 0.000 |

## Supplementary materials

**Table S1.** Full seroprevalence results by site over time for 4 assays

*See accompanying spreadsheet*, **seroprevalence 2020-12-15.xlsx**

**Figure S2.**
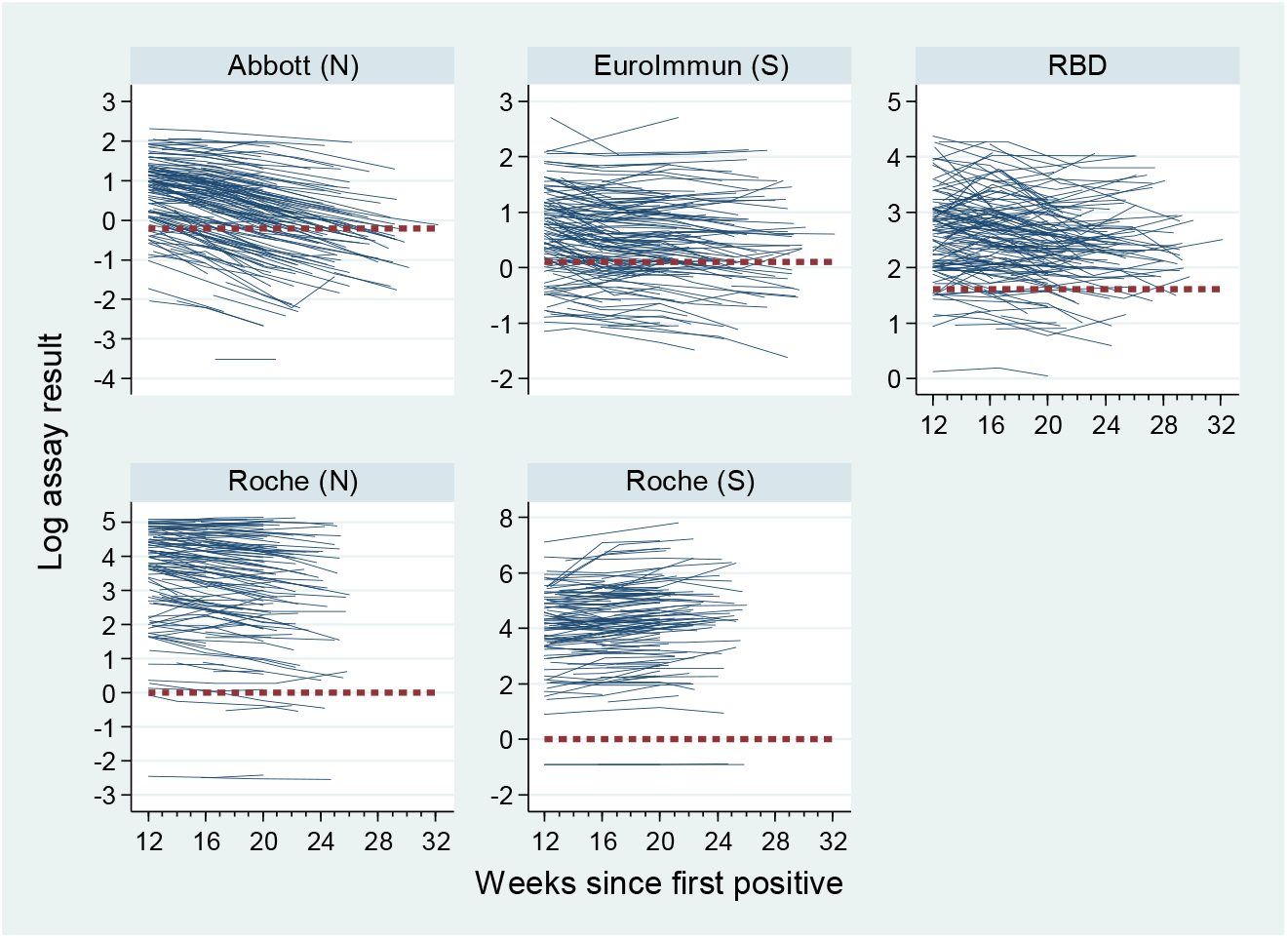
Individual responses from 12 weeks onwards using five different assays.

**Table S3.**
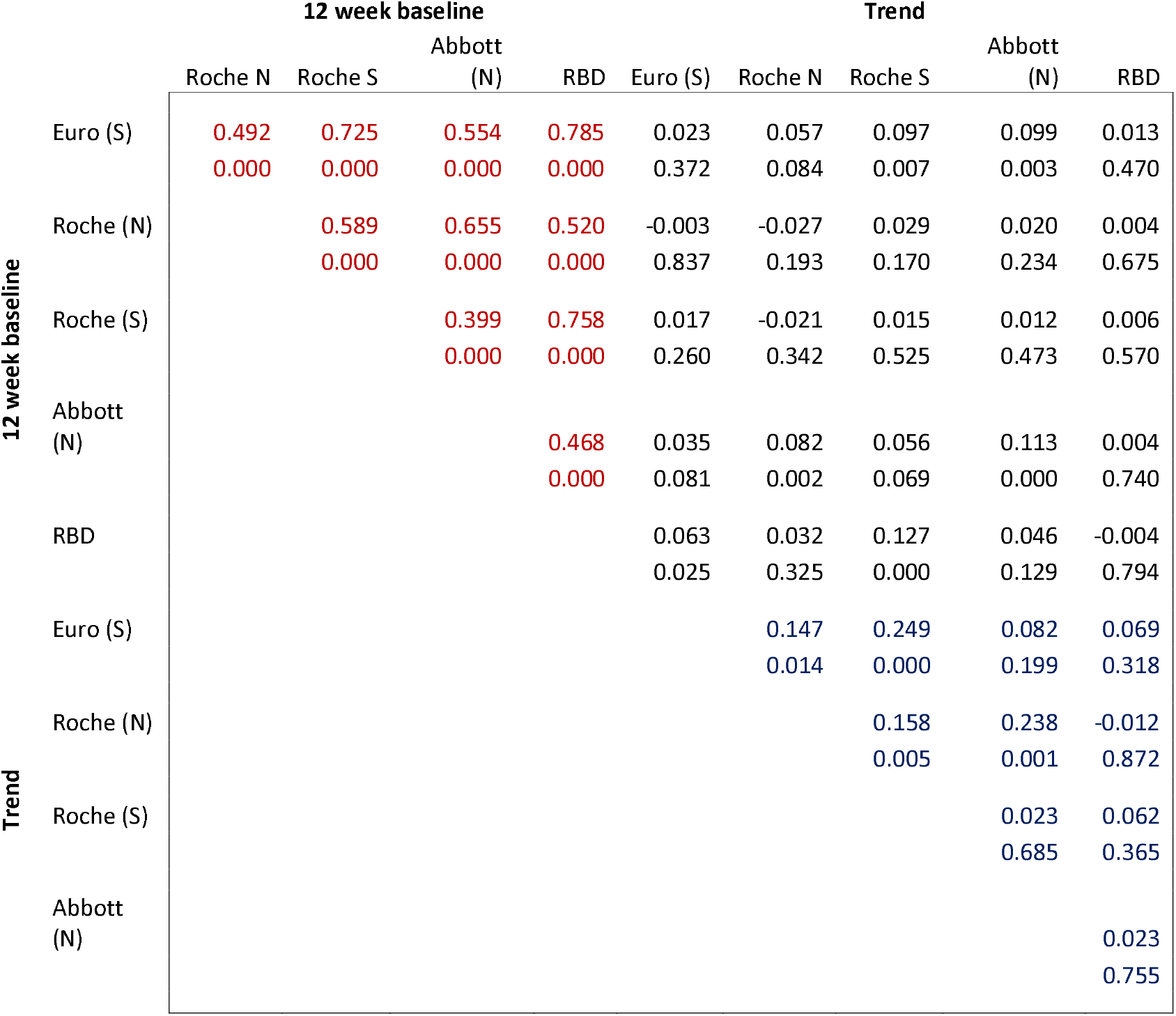
Correlations between individual-level random effects (intercept and trend) between different assays; each cell shows the posterior median of the correlation coefficient and 2-sided Bayesian p-value for the coefficient being zero. Red cells indicate correlations in intercepts, blue cells correlations in trends, and black cells intercept-trend correlations.

**Figure S4.**
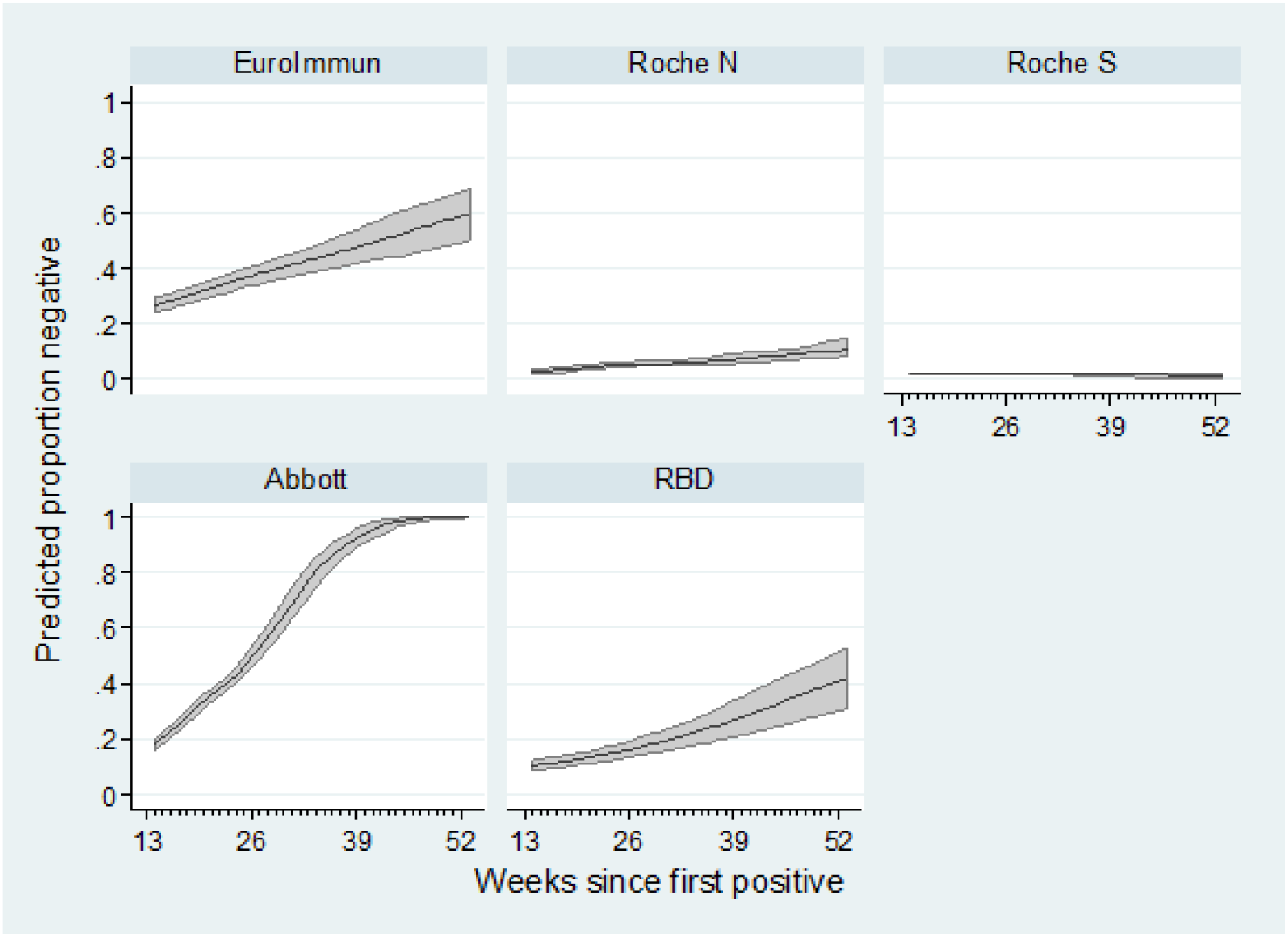
Projected proportions reverting to negative test status over time, from 13 weeks after the first positive test, under five different assays. Posterior medians and 95% credible intervals from Bayesian random effects model.

**Figure S5.**
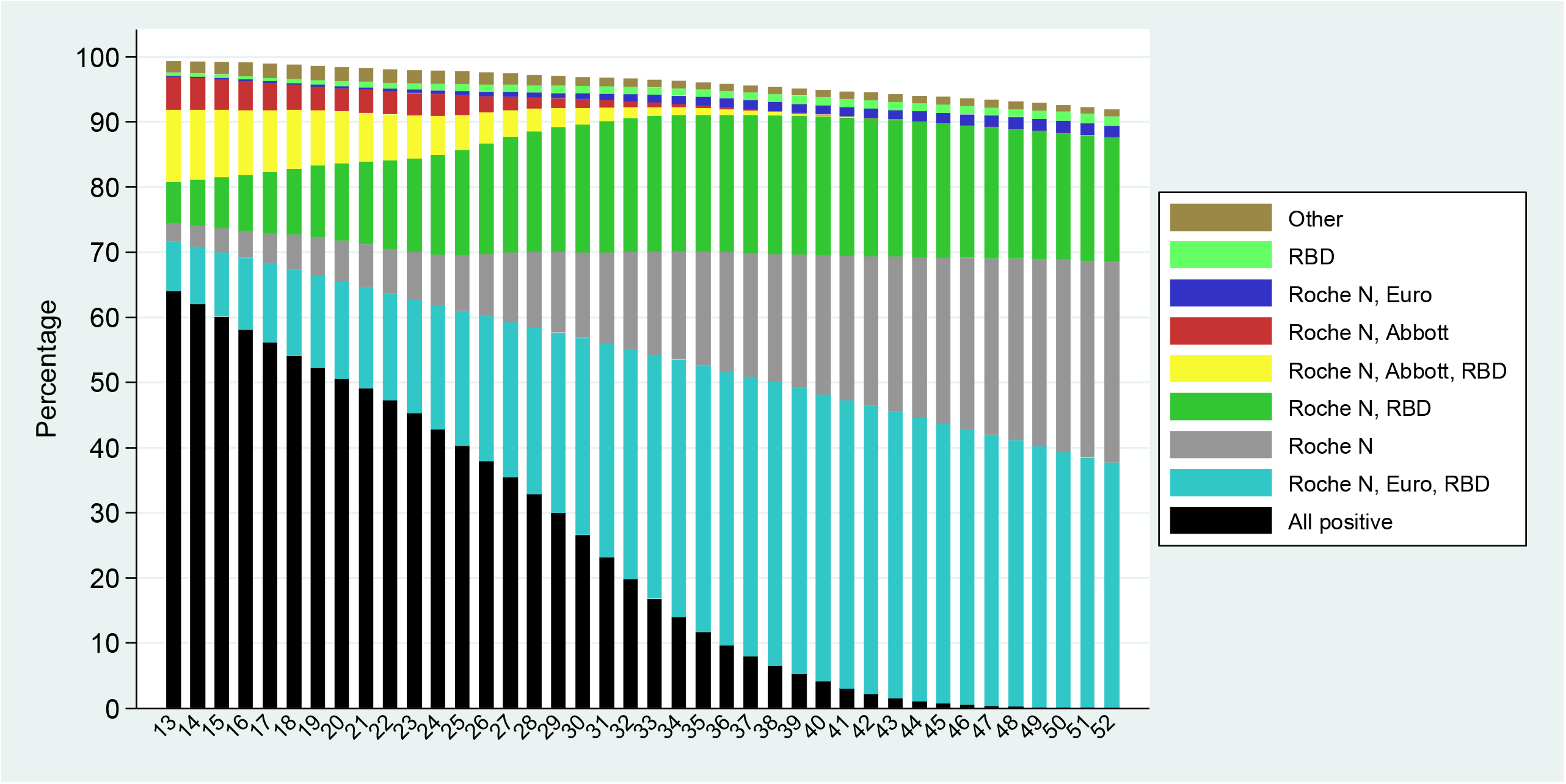
Predicted multivariate assay status by week, from 13 weeks after the first positive test; mean proportions from Bayesian random effects model. Nearly all are predicted to remain positive under Roche S, therefore EuroImmun, Roche N, Abbott and RBD are examined. There are 16 possible combinations of positive/negative for the 4 assays; combinations with <1% are grouped as “other”.

## Notes

### Competing Interest Statement

The authors have declared no competing interest.

### Funding Statement

This research was internally funded by Public Health England. BP is supported by the NIHR Manchester Biomedical Research Centre. At no time did any other author or their institutions receive payment or services from a third party for any aspect of the submitted work.

### Author Declarations

This study was approved by PHE Research Support and Governance Office (R&D REGG Ref NR 0190)

### Summary of Updates

The analysis has been updated to include a longer follow-up time (6 months, compared to 3 months previously) and now looks at 5 different assays (was 2). Initial antibody responses are modelled (first 12 weeks), and subsequent trends after 12 weeks. Predicted times to negativity are produced and agreement between assays assessed. The overall conclusion is that waning slows after 12 weeks, but is still present; although some assays continue to exhibit a rapid decline.

## References

1. He J, Guo Y, Mao R, Zhang J. Proportion of asymptomatic coronavirus disease 2019: A systematic review and meta-analysis. J Med Virol. 2020 Jul 21; Available from: http://www.ncbi.nlm.nih.gov/pubmed/32691881

2. Wu X, Liu L, Jiao J, Yang L, Zhu B, Li X. Characterisation of clinical, laboratory and imaging factors related to mild vs. severe covid-19 infection: a systematic review and meta-analysis. Ann Med. 2020 Aug 11;1–11. Available from: http://www.ncbi.nlm.nih.gov/pubmed/32755287

3. Kumar A, Arora A, Sharma P, Anikhindi SA, Bansal N, Singla V, et al. Clinical Features of COVID-19 and Factors Associated with Severe Clinical Course: A Systematic Review and Meta-Analysis. SSRN. 2020 Apr 21;3566166. Available from: http://www.ncbi.nlm.nih.gov/pubmed/32714109

4. Iversen K, Bundgaard H, Hasselbalch RB, Kristensen JH, Nielsen PB, Pries-Heje M, et al. Risk of COVID-19 in health-care workers in Denmark: an observational cohort study. Lancet Infect Dis. 2020 Aug 3;0(0). Available from: http://www.ncbi.nlm.nih.gov/pubmed/32758438

5. Watson J, Whiting PF, Brush JE. Interpreting a covid-19 test result. BMJ. 2020;369(May):1–7. Available from: http://dx.doi.org/doi:10.1136/bmj.m1808

6. National COVID-19 surveillance reports - GOV.UK. Available from: https://www.gov.uk/government/publications/national-covid-19-surveillance-reports

7. Koopmans M, Haagmans B. Assessing the extent of SARS-CoV-2 circulation through serological studies. Nat Med. 2020;26(8):1171–2. Available from: http://www.ncbi.nlm.nih.gov/pubmed/32719488

8. Tanne JH. Covid-19: US cases are greatly underestimated, seroprevalence studies suggest. BMJ. 2020 Jul 24;370:m2988. Available from: http://www.ncbi.nlm.nih.gov/pubmed/32709608

9. Addetia A, Crawford KH, Dingens A, Zhu H, Roychoudhury P, Huang M-L, et al. Neutralizing antibodies correlate with protection from SARS-CoV-2 in humans during a fishery vessel outbreak with high attack rate. medRxiv Prepr Serv Heal Sci. 2020 Aug 14; Available from: http://www.ncbi.nlm.nih.gov/pubmed/32817980

10. Kampen JJA van, Vijver DAMC van de, Fraaij PLA, Haagmans BL, Lamers MM, Okba N, et al. Shedding of infectious virus in hospitalized patients with coronavirus disease-2019 (COVID-19): duration and key determinants. medRxiv. 2020 Jun 9;2020.06.08.20125310. Available from: https://www.medrxiv.org/content/10.1101/2020.06.08.20125310v1

11. Okell LC, Verity R, Watson OJ, Mishra S, Walker P, Whittaker C, et al. Have deaths from COVID-19 in Europe plateaued due to herd immunity? Lancet (London, England). 2020;395(10241):e110–1. Available from: http://www.ncbi.nlm.nih.gov/pubmed/32534627

12. Grandjean L, Saso A, Ortiz A, Lam T, Hatcher J, Thistlethwaite R, et al. Humoral Response Dynamics Following Infection with SARS-CoV-2. medRxiv. 2020 Jul 22;2020.07.16.20155663. Available from: https://www.medrxiv.org/content/10.1101/2020.07.16.20155663v2

13. Liu A, Li Y, Peng J, Huang Y, Xu D. Antibody responses against SARS-CoV-2 in COVID-19 patients. J Med Virol. 2020 Jun 30; Available from: http://www.ncbi.nlm.nih.gov/pubmed/32603501

14. Patel MM, Thornburg NJ, Stubblefield WB, Talbot HK, Coughlin MM, Feldstein LR, et al. Change in Antibodies to SARS-CoV-2 Over 60 Days Among Health Care Personnel in Nashville, Tennessee. JAMA. 2020 Sep 17; Available from: https://jamanetwork.com/journals/jama/fullarticle/2770928

15. Kissler SM, Tedijanto C, Goldstein E, Grad YH, Lipsitch M. Projecting the transmission dynamics of SARS-CoV-2 through the postpandemic period. Science (80-). 2020 May 22;368(6493):860–8. Available from: http://www.ncbi.nlm.nih.gov/pubmed/32291278

16. Prime Minister’s statement on coronavirus (COVID-19): 23 March 2020 - GOV.UK. Available from: https://www.gov.uk/government/speeches/pm-address-to-the-nation-on-coronavirus-23-march-2020

17. Coronavirus (COVID-19) Infection Survey pilot - Office for National Statistics. Available from: https://www.ons.gov.uk/peoplepopulationandcommunity/healthandsocialcare/conditionsanddiseases/bulletins/coronaviruscovid19infectionsurveypilot/englandandwales14august2020

18. Amirthalingam G, Whitaker H, Brooks T, Brown K, Hoschler K, Linley E, et al. Evaluating the seroprevalence of SARS-CoV 2 amongst adult blood donors in London and changes following the introduction of Public Health and social measures. Emerg Infect Dis. 2020;submitted.

19. Edridge AWD, Kaczorowska J, Hoste ACR, Bakker M, Klein M, Loens K, et al. Seasonal coronavirus protective immunity is short-lasting. Nat Med. 2020 Sep 14;1–3. Available from: http://www.nature.com/articles/s41591-020-1083-1

20. Long Q-X, Liu B-Z, Deng H-J, Wu G-C, Deng K, Chen Y-K, et al. Antibody responses to SARS-CoV-2 in patients with COVID-19. Nat Med. 2020;26(6):845–8. Available from: http://www.ncbi.nlm.nih.gov/pubmed/32350462

21. Zhao J, Yuan Q, Wang H, Liu W, Liao X, Su Y, et al. Antibody responses to SARS-CoV-2 in patients of novel coronavirus disease 2019. Clin Infect Dis. 2020 Mar 28; Available from: http://www.ncbi.nlm.nih.gov/pubmed/32221519

22. Wang X, Guo X, Xin Q, Pan Y, Li J, Chu Y, et al. Neutralizing Antibodies Responses to SARS-CoV-2 in COVID-19 Inpatients and Convalescent Patients. medRxiv. 2020 Apr 23;2020.04.15.20065623. Available from: https://www.medrxiv.org/content/10.1101/2020.04.15.20065623v3

23. Seow J, Graham C, Merrick B, Acors S, Steel KJA, Hemmings O, et al. Longitudinal evaluation and decline of antibody responses in SARS-CoV-2 infection. medRxiv. 2020 Jul 11;2020.07.09.20148429. Available from: https://www.medrxiv.org/content/10.1101/2020.07.09.20148429v1

24. Long Q-X, Tang X-J, Shi Q-L, Li Q, Deng H-J, Yuan J, et al. Clinical and immunological assessment of asymptomatic SARS-CoV-2 infections. Nat Med. 2020 Aug 18;26(8):1200–4. Available from: http://www.nature.com/articles/s41591-020-0965-6

25. Ibarrondo FJ, Fulcher JA, Goodman-Meza D, Elliott J, Hofmann C, Hausner MA, et al. Rapid Decay of Anti-SARS-CoV-2 Antibodies in Persons with Mild Covid-19. N Engl J Med. 2020;383(11):1085–7. Available from: http://www.ncbi.nlm.nih.gov/pubmed/32706954

26. Gudbjartsson DF, Norddahl GL, Melsted P, Gunnarsdottir K, Holm H, Eythorsson E, et al. Humoral Immune Response to SARS-CoV-2 in Iceland. N Engl J Med. 2020 Sep 1;NEJMoa2026116. Available from: http://www.nejm.org/doi/10.1056/NEJMoa2026116

27. Guihot A, Litvinova E, Autran B, Debré P, Vieillard V. Cell-Mediated Immune Responses to COVID-19 Infection. Front Immunol. 2020;11:1662. Available from: http://www.ncbi.nlm.nih.gov/pubmed/32719687

28. Grifoni A, Weiskopf D, Ramirez SI, Mateus J, Dan JM, Moderbacher CR, et al. Targets of T Cell Responses to SARS-CoV-2 Coronavirus in Humans with COVID-19 Disease and Unexposed Individuals. Cell. 2020;181(7):1489–1501.e15. Available from: http://www.ncbi.nlm.nih.gov/pubmed/32473127

29. Le Bert N, Tan AT, Kunasegaran K, Tham CYL, Hafezi M, Chia A, et al. SARS-CoV-2-specific T cell immunity in cases of COVID-19 and SARS, and uninfected controls. Nature. 2020 Aug 20;584(7821):457–62. Available from: http://www.nature.com/articles/s41586-020-2550-z

30. Chen Z, John Wherry E. T cell responses in patients with COVID-19. Nat Rev Immunol. 2020 Sep 29;20(9):529–36. Available from: http://www.nature.com/articles/s41577-020-0402-6

31. Zuo J, Dowell A, Pearce H, Verma K, Hm L, Begum J, et al. Robust SARS-CoV-2-specific T-cell immunity is maintained at 6 months following primary infection. bioRxiv. 2020; Available from: https://doi.org/10.1101/2020.11.01.362319

32. Wu L-P, Wang N-C, Chang Y-H, Tian X-Y, Na D-Y, Zhang L-Y, et al. Duration of antibody responses after severe acute respiratory syndrome. Emerg Infect Dis. 2007 Oct;13(10):1562– 4. Available from: http://www.ncbi.nlm.nih.gov/pubmed/18258008

33. Payne DC, Iblan I, Rha B, Alqasrawi S, Haddadin A, Al Nsour M, et al. Persistence of Antibodies against Middle East Respiratory Syndrome Coronavirus. Emerg Infect Dis. 2016;22(10):1824– 6. Available from: http://www.ncbi.nlm.nih.gov/pubmed/27332149

